# Enteric pathogen infections among infants in rural Bangladesh: prevalence, trial impact, and associations with enteric dysfunction and growth

**DOI:** 10.64898/2026.06.24.26356432

**Authors:** Anna A. Müller-Hauser, Nathalie J. Lambrecht, Shafinaz Sobhan, Jillian L. Waid, Tarique Md. Nurul Huda, Dennis Nurjadi, Amanda S. Wendt, Mahbubur Rahman, Sabine Gabrysch

## Abstract

**Background:** Repeated enteric infections and chronic enteric dysfunction have been associated with undernutrition in children. Interventions that reduce enteric pathogen exposure in young children could thereby improve growth and developmental outcomes. We assessed enteric pathogen prevalence in Bangladeshi infants, the impact of a combined homestead food production and food hygiene intervention on pathogen infections, and associations between pathogen infections, enteric dysfunction markers, and child growth outcomes.

**Methods:** We analyzed panel data from 231 children born between April and December 2018 within the Food and Agricultural Approaches to Reducing Malnutrition (FAARM) cluster-randomized trial in Sylhet, Bangladesh. Stool samples were collected at 0–3, 6–8, and 10–13 months of age and assessed for enteric dysfunction biomarkers (myeloperoxidase, alpha-1 antitrypsin, and neopterin) by ELISA and 14 enteric pathogens by multiplex RT-PCR. Diarrhea prevalence was recorded using 7-day recall. Child length and weight were measured at birth and trial endline. Multilevel regression assessed the intervention effect and quantified associations between pathogen exposure, enteric dysfunction, and growth outcomes.

**Findings:** Enteric pathogen prevalence was high (84%) despite low 7-day diarrhea prevalence (5%), and co-infections were common. There was no intervention effect on the prevalence of enteric pathogens. *Shigella* spp. and *Giardia lamblia* were associated with higher myeloperoxidase concentrations, while sapovirus was associated with higher alpha-1 antitrypsin concentrations. Repeated protozoan infections (mainly *Giardia lamblia*) were associated with lower length-for-age, while repeated viral infections (mainly rotavirus and sapovirus) and *Cryptosporidium* infections were associated with lower weight-for-height and weight-for-age. There was marginal evidence that bacterial infections were associated with lower length-for-age.

**Conclusion:** The intervention was insufficient to reduce the high burden of enteric pathogens in infants, and subclinical infections were associated with enteric dysfunction and poorer growth outcomes. Comprehensive strategies addressing all key exposure pathways are needed to limit pathogen infections and their consequences for child development.

**Author Summary:** Infections of the gut are extremely common in young children in low-income settings and can cause lasting harm even when they do not produce obvious symptoms like diarrhea. In this study, we examined stool samples from infants in rural Bangladesh and found that over 80% carried at least one enteric pathogen at any given time, despite only 5% showing signs of diarrhea. This confirms that the majority of these infections go unnoticed by caregivers and standard health monitoring.

We investigated whether a combined program promoting home gardens, poultry rearing, better child feeding, and improved household food hygiene could reduce these infections, but found no measurable effect on pathogen burden. We found clear links between infection with specific pathogens and both gut inflammation and poorer child growth: protozoan parasites were associated with stunting, while viruses and Cryptosporidium were linked to wasting and underweight.

These findings underscore that visible illness is a poor proxy for pathogen exposure in infants in low-income settings, and that more comprehensive strategies, such as transformative water, sanitation, and hygiene approaches, may be needed to meaningfully reduce the hidden burden of enteric infections and their consequences for child development.

## Introduction

Undernutrition is a leading risk factor for death, poor health outcomes, and developmental deficits in children, particularly affecting children in low-and middle-income countries (LMIC) (1, 2). Recurrent enteric infections with or without symptomatic diarrheal disease, as well as inadequate dietary intake, contribute to undernutrition (3–7). While the burden of diarrhea is high among young children in LMIC (6, 8), asymptomatic enteric pathogen infections are even more widespread and thought to contribute to environmental enteric dysfunction (EED), a subclinical intestinal inflammatory disorder (9–14). EED is characterized by increased intestinal permeability, reduced nutrient absorption, and intestinal and systemic immune activation (15, 16). Both EED and asymptomatic enteric pathogen infections have been associated with undernutrition and resulting growth deficits in children, though with varying strength of associations, depending on the study setting, age of children, anthropometric measures taken, as well as biomarkers of EED and pathogens studied (9, 11, 12, 17–22). Nevertheless, these results suggest that interventions that successfully reduce enteric pathogen exposure in young children reduce EED and improve growth outcomes in early childhood.

Recent studies evaluating interventions aimed at improving water, sanitation, and hygiene (WASH) infrastructure have shown mixed success in reducing enteric infections, diarrhea, and child growth deficits. While a sanitation intervention in Mozambique had no effect on diarrhea prevalence or child growth (23), a water chlorination intervention in Bangladesh reduced diarrhea prevalence (24). Two sanitation interventions in Mali and India improved child growth but without a reduction in diarrhea prevalence (25, 26). Three large factorial trials studying the effects of combined and separate nutrition and WASH interventions showed mixed results on diarrhea, enteric pathogen infections, and growth in children. While all three trials in Zimbabwe, Bangladesh, and Kenya saw a modest intervention effect on child growth outcomes (27–29), only the study in Bangladesh reduced diarrhea prevalence (28). In Zimbabwe, the intervention reduced the number of parasites detected, but not bacterial or viral pathogens (30). In all three trials, there was no WASH intervention effect on child growth outcomes, and impacts on growth were observed only in trial arms that also received a nutrition intervention, suggesting that the WASH interventions did not interrupt relevant contamination pathways to sufficiently reduce enteric pathogen exposure (31). Therefore, a better understanding of the pathogens and pathways contributing to undernutrition in LMIC settings could help to inform intervention strategies.

Nested within the *Food and Agricultural Approaches to Reducing Malnutrition* (FAARM) cluster-randomized controlled trial, the *Food Hygiene to reduce Environmental Enteric Dysfunction* (FHEED) study was designed to assess the effect of a combined homestead food production and food hygiene intervention on enteric pathogen infection and EED. The intervention effect on EED has already been analyzed in the full FHEED sample cross-sectionally (32). In the present study, we assess the intervention effect on enteric pathogen infections in a cohort of FHEED children followed over their first year of life. We also determine the overall prevalence of enteric pathogen infections among these infants and examine the associations between enteric pathogen infections, markers of EED, and growth outcomes.

## Methods

### Study design and participants

We used data from the FAARM cluster-randomized trial (ClinicalTrials.gov, ID: NCT02505711) conducted from 2015 to 2020, and the FHEED sub-study. The FAARM trial evaluated the impact of a homestead food production (HFP) program on child undernutrition, with height-for-age as the primary outcome. The intervention was implemented in rural Habiganj district, Sylhet division, Bangladesh, by the international non-governmental organization *Helen Keller International* from mid-2015 to late-2018. The FAARM trial enrolled 2705 married women in 96 settlements (geographic clusters) assigning the settlements to intervention and control conditions (48 in each arm) using covariate-constrained randomization. The HFP program included training on year-round gardening, poultry rearing, and improved nutrition and hygiene practices (33). As part of the FHEED sub-study, an additional food hygiene component was implemented in the third year of the intervention (from June 2017 to February 2018) to strengthen household food hygiene practices around food preparation and child feeding, using a behavior-centered design and emotional drivers (34). The FHEED sub-study was designed to analyze the effect of the combined HFP and food hygiene intervention on household food hygiene practices, complementary food contamination, enteric pathogen infection and EED in young children, and how all of these factors eventually influence child growth.

### Data collection

FHEED collected data from a birth cohort of children born between April and December 2018, which were followed over their first year of life, with surveys every two months until the FAARM endline survey, conducted from September to December 2019. The analysis presented here is based on three rounds of data and samples collected from the 231 cohort children at ages 0-3 months, 6-8 months, and 10-13 months (Figure 1).

**Figure 1:**
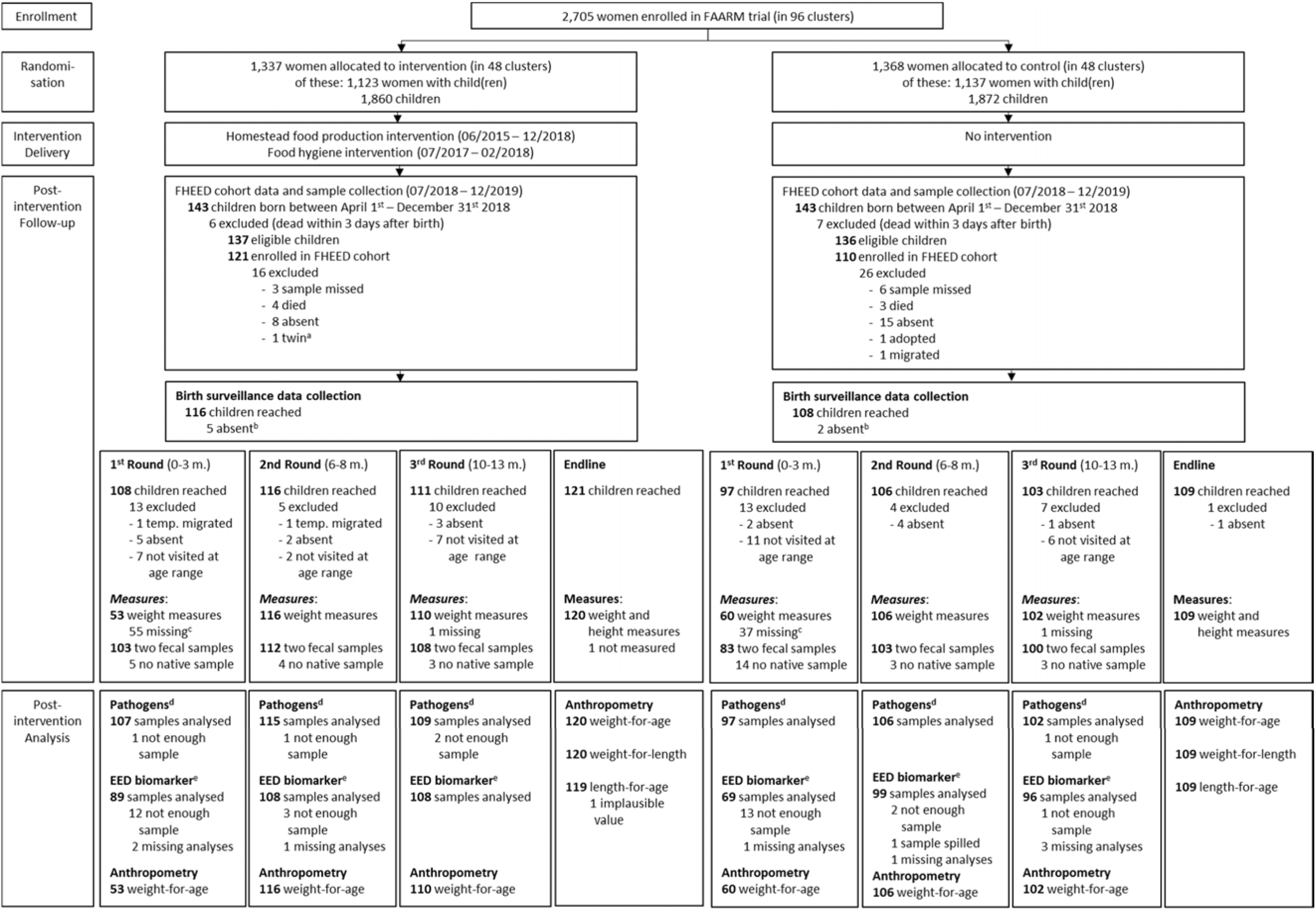
Study flow diagram. ^a^ in case of twins, only one of the two children were enrolled in the FHEED cohort; ^b^ children with birth surveillance measures after 28 days post birth were considered absent for birth surveillance. For analyses, measures were replaced by the average anthropometry measures at birth of the FHEED cohort.; ^c^ children sampled in the 1^st^ round of FHEED cohort surveys were not weighed, weight measurements were conducted from the 2^nd^ cohort round onwards; ^d^ conducted from buffered fecal samples; ^e^ conducted from native fecal samples. Abbreviations: FAARM: Food and Agricultural Approaches to Reducing Malnutrition; FHEED: Food Hygiene to reduce Environmental Enteric Dysfunction; 0-3m.: 0-3 months of age, 6-8m.: 6-8 months of age, 10-13m.: 10-13 months of age; temp. migrated: temporarily migrated

We used data from the following FAARM and FHEED surveys: 1) characteristics of mothers and households from the FAARM baseline survey (March to May 2015), 2) child date of birth, sex, and anthropometric measures at birth from the FAARM birth surveillance (April to December 2018), 3) child health status, weight, season of sampling, and fecal samples collected in 7 FHEED cohort surveys (July 2018 to December 2019), and 4) child health status and anthropometric measures from the FAARM endline survey (September to December 2019).

The FAARM baseline survey collected data on mother and household characteristics, such as mothers’ age, education, and height, and household religion, wealth, and sanitation facilities. We used household asset information to estimate each household’s position within the 2014 Demographic and Health Survey national wealth quintiles according to Equity Tool guidelines (35, 36). Mother’s education was measured as the number of school years completed and grouped into none, some or completed primary education, and some secondary education and higher. A handwashing station was considered functional when it was equipped with water, a cleaning agent, and a pouring device. Access to basic sanitation facilities was defined as the presence of a flush or pour flush toilet connected to a piped sewer system, septic tank or pit latrine, a pit latrine with a slab (including ventilated pit latrine), and a composting toilet, not shared with other households (37).

Data on recent child illness were collected alongside fecal sampling, both during FHEED cohort surveys and at endline. Caregivers were asked if their child was sick with loose or mushy stools within the 7 days prior to the survey; if caregivers reported loose or mushy stools, they were asked for the number of stools passed on the worst day. A child was considered sick with diarrhea when a caregiver reported three or more loose or mushy stools on at least one day within 7 days prior to the survey. Season of sampling was defined according to the six seasons in Bangladesh, each lasting 2 months, beginning in mid-April.

Child anthropometric measures (length and weight) were taken at birth and at the FAARM endline survey by trained data collectors in line with FANTA guidelines (38). For anthropometry at birth, we aimed to visit mothers and newborns within 72 hours of birth, and for children measured after 28 days post birth, birth surveillance measures were considered missing and replaced by the average anthropometry measures at birth of the FHEED cohort. Two independent measurements were taken for each parameter, and the results were averaged; in case the results were not in alignment, a third measure was taken and the data collector indicated the best measure. Between birth and endline, we also measured child weight at each FHEED cohort survey. Length-for-age, weight-for-length, and weight-for-age z-scores were calculated using WHO reference standards (39).

Questionnaires were administered by trained data collection officers who conducted face-to-face interviews. All survey data were collected using tablet-based Open Data Kit (ODK) software (40).

### Biological sample collection

Fecal samples of children were collected by mothers or other caregivers. They were provided with two sterile stool collection tubes (one without additives and one with 7 mL RNAlater®, Sigma-Aldrich, USA) in a Ziplock bag, gloves, and instructions on how to collect the stool sample from their children (infographic as well as verbal explanation and practical demonstration by the field team). Mothers were asked to first fill the container without additives, and then to transfer one small spoon of fecal material (∼ 1 g of stool) to the container with RNAlater® and shake by hand. If possible, mothers collected the child’s first stool sample in the morning and stored the collection tubes in the Ziplock bag in a shady spot. The field team collected the stool samples from the households in the morning and stored them in a cool box with ice packs, monitoring the temperature in the cool box (8-10°C) at all times. In case a child did not pass stool in the morning, the sample collector returned to the household later that day to collect the sample. Samples were brought to the field laboratory by noon. First, native samples (without additives) were aliquoted, then the buffered samples (in RNAlater®) were mixed thoroughly by vortexing at full speed, and aliquoted (1.8 mL stool-buffer mix per tube). All samples were stored at −20°C until shipment to Germany, where they were stored at −80°C until further analyses.

### Biomarker analyses

Biomarkers of EED (myeloperoxidase [MPO] and neopterin [NEO], measuring intestinal inflammation, and alpha-1 antitrypsin [AAT], measuring intestinal permeability) were assessed in native stool samples by enzyme-linked immunosorbent assay (ELISA). Commercially available ELISA kits for MPO (R&D Systems, USA), NEO (IBL International, Germany), and AAT (R&D Systems, USA) were used according to the manufacturers’ instructions. A detailed description of the assay procedures has been published elsewhere (32). In brief, about 100 mg of fecal sample were homogenized in 1 mL ice-cold sample diluent (Ref. KENO751, IBL International, Germany) for 2 x 1 minute using a bead beater and centrifuged at 2500 x g at 4°C to clear the sample. Supernatants were either used immediately in ELISAs or frozen at −20°C for short-term storage (used within 5 days). For analysis, supernatants were diluted 1:5000 (MPO), 1:20000 (AAT), and 1:100 (NEO) with the respective assay buffer. ELISAs were run in technical duplicates. Samples with out-of-range values were re-tested at an appropriate dilution.

### Pathogen analyses

Microbial nucleic acid was extracted from buffered stool samples using the AllPrep® PowerFecal® DNA/RNA Kit (Qiagen, Germany). We followed the manufacturer’s instructions, with slight alterations to the protocol: stool samples stored in RNAlater® (∼250 mg of stool) were thawed on ice, centrifuged for 5 minutes at 13,000 x g at 4°C, and washed 3 times with 1 mL ice-cold Phosphate-Buffered-Saline, to ensure full removal of RNAlater®. Lysis and homogenization were done according to the protocol, with an additional Phenol/Chloroform step, following RNA and DNA purification steps. RNA was eluted in 60 µL RNase-free water (Qiagen, Germany), and RNA concentration was measured using a NanoDrop^TM^ 2000 spectrophotometer (ThermoFisher Scientific, USA). DNA was eluted in 70 µL Elution Buffer (Qiagen, Germany), and DNA concentration was measured using a Qubit^TM^ Flex Fluorometer (Invitrogen^TM^, USA). RNA and DNA samples were stored at −80°C until further analyses.

For enteric pathogen analysis, the following commercially available diagnostic test kits, based on real-time polymerase chain reaction (RT-PCR), were used according to manufacturers’ instructions: FTD^TM^ Bacterial gastroenteritis (Siemens Healthineers, Germany) for detection of the bacterial enteric pathogens *Campylobacter jejuni/coli/lari*, toxin producing *Clostridioides difficile*, *Salmonella enterica*, *Shigella dysenterica/flexneri/boydii/sonnei* and enteroinvasive *Escherichia coli* (EIEC), verotoxin-producing *Escherichia coli* (VTEC), and *Yersinia enterocolitica*; FTD^TM^ Viral gastroenteritis (Siemens Healthineers, Germany) for detection of the viral enteric pathogens norovirus genogroup (G)I and GII, rotavirus, human astrovirus, human adenovirus, and sapovirus; FTD^TM^ Stool parasites (Siemens Healthineers, Germany) for detection of the protozoan parasites *Cryptosporidium* spp., *Entamoeba histolytica*, and *Giardia lamblia*. Samples were run on 96-well plates, with each plate also containing a positive and negative control, using the QuantStudio™ 5 Real-Time PCR System. Each amplification well contained 12.5 µL 2xRT-PCR Buffer, 1.5 µL Primer/Probe Mix, 1 µL RT-PCR-Enzyme, and 10 µL sample (or 10 µL positive control or negative control). Samples went through the following PCR program: 15 minutes at 50°C, 1 minute at 94°C, 40 cycles of 8 seconds at 94°C followed by 1 minute at 60°C. Threshold cycles (C_t_) were determined using the VERSANT® kPCR MiPLX AD software (Siemens Healthineers, Germany). A C_t_ cut-off of ≤ 35 amplification cycles was applied to define positive detection of the respective pathogen. In case a positive control did not amplify or a negative control did amplify, the run was considered invalid.

### Statistical analysis

All data analyses were performed in Stata SE version 14.2 (StataCorp, College Station, TX, USA). Socio-demographic characteristics of participants and their households at baseline, disaggregated by intervention group, were summarized using arithmetic means and standard deviations for continuous variables and proportions for categorical variables.

For statistical analyses, EED biomarker concentrations were log-transformed to approximate normal distribution. We calculated Pearson correlations between concentrations of each biomarker with the other two, stratified by child age (0-3, 6-8, and 10-13 months). Predicted concentrations of EED biomarkers, predicted diarrhea prevalence, and predicted pathogen prevalence by intervention allocation and by child age were calculated using the *margins* command after multilevel mixed-effects linear or logistic regression models, adjusting for season of sampling and sex of the child, with settlement- and child-level random effects. Intervention effects on pathogen prevalence were assessed using mixed-effects linear or logistic regression models, adjusting for season of sampling and age and sex of the child, with settlement-level random effects. Intervention effects on EED biomarkers, diarrhea prevalence, and pathogen prevalence by child age were calculated using an interaction term between intervention allocation and child age group. Predicted biomarker concentrations and pathogen prevalences were calculated using the *margins* command, and the *lincom* command was used to calculate point estimates and 95% confidence intervals. For some pathogen outcomes, mixed-effects logistic regression models did not converge; in that case, we used cluster-robust standard errors with clustering at settlement level.

To assess the associations between pathogen exposure and biomarkers of EED and pathogen exposure and weight-for-age z-score (at the time of the FHEED cohort surveys), we employed mixed-effects linear regression models with settlement- and child-level random effects. Pathogen exposure was modeled as the presence of pathogen groups (presence of any bacteria, any virus, or any parasite), or the presence of single pathogens, adjusting for the presence of other pathogens, respectively. Models were further adjusted for season of sampling, intervention allocation, age and sex of the child, and, for the models with weight-for-age z-score as the outcome, we additionally adjusted for weight-for-age z-score at birth and maternal height.

Similarly, the associations between pathogen exposure or biomarkers of EED and i) length-for-age, ii) weight-for-age, or iii) weight-for-height z-score at study endline was assessed using mixed-effects linear regression models, adjusting for the presence of other pathogens or concentration of other EED biomarkers, respectively. Models were further adjusted for age and sex of the child, length-for-age, weight-for-age or weight-for-height z-score at birth, respectively, maternal height, and intervention allocation, with settlement-level random effects. Cumulative pathogen exposure was modeled as the number of stools a) positive for a pathogen group or b) positive for a single pathogen (out of the maximum three stools obtained), these models were additionally adjusted for the number of rounds a stool sample was obtained. Single EED biomarkers were modeled as the average concentration of the log-transformed biomarkers, obtained from three rounds of stool collection. All model specifications are detailed in Suppl. Table 1.

### Ethical considerations

The FAARM trial protocol was positively reviewed by ethics boards at Heidelberg University’s Medical Faculty in Germany (Ref.: S-121/2014) and the James P. Grant School of Public Health, BRAC University in Bangladesh (Ref.: 37A), and the FHEED study protocol by Heidelberg University’s Medical Faculty (Ref.: S-606/2017) and icddr,b in Bangladesh (Ref.: PR-17126). Informed written consent by signature or thumbprint was obtained from the primary caregivers of the enrolled children.

## Results

This analysis includes data from 231 children enrolled in the FHEED birth cohort (85% of eligible children born between April 1^st^, 2018 and December 31^st^, 2018). Survey data and stool samples were collected at child ages 0-3, 6-8, and 10-13 months, with 183 children (79%) providing data and samples at all three time points, 44 children (19%) at two, and 4 children (2%) at only one (Table 1). Child height and weight at study endline in 2019 were measured in 228 of 231 children (99%). Overall, 641 observations of 231 children were analyzed. Figure 1 provides the number of children included in the analysis, as well as reasons for exclusion.

**Table 1:** Baseline characteristics of households, mothers, and children of the FHEED cohort in rural Sylhet, Bangladesh.

| Characteristics |  | Overall<br>n=231 | Control<br>n=110 | Intervention<br>n=121 |
| --- | --- | --- | --- | --- |
| <u>Household characteristics</u> |  |  |  |  |
| Household wealth <sup>a</sup> | Least wealthy (%) | 12 | 12 | 12 |
|  | Lower (%) | 29 | 34 | 25 |
|  | Middle (%) | 26 | 26 | 26 |
|  | Upper (%) | 27 | 24 | 29 |
|  | Wealthiest (%) | 6 | 4 | 8 |
| Religion | Muslim (%) | 74 | 70 | 77 |
|  | Hindu (%) | 26 | 30 | 23 |
| Number of household members |  | 7.4 (3.4) | 7.4 (3.5) | 7.4 (3.4) |
| Improved water source <sup>b</sup> (%) |  | 97 | 95 | 98 |
| Functional handwashing station <sup>c</sup> (%) |  | 55 | 49 | 60 |
| Access to basic latrines <sup>d</sup> (%) |  | 39 | 32 | 45 |
| <u>Mother's characteristics</u> |  |  |  |  |
| Mother's education <sup>e</sup> | None (%) | 15 | 15 | 16 |
|  | Partial/complete primary (%) | 48 | 49 | 46 |
|  | At least partial secondary (%) | 37 | 36 | 38 |
| Mother's age at baseline (years) |  | 22.6 (3.6) | 22.6 (3.7) | 22.6 (3.5) |
| <u>Child characteristics</u> |  |  |  |  |
| Sex | Male (%) | 52 | 56 | 48 |
|  | Female (%) | 48 | 44 | 52 |
| Number of fecal samples | One (%) | 2 | 2 | 2 |
|  | Two (%) | 19 | 18 | 20 |

|  | Three (%) | 79 | 80 | 78 |
| --- | --- | --- | --- | --- |
| Age in months round 1, n=205 |  | 1.4 (0.95) | 1.4 (0.96) | 1.4 (0.96) |
| Age in months round 2, n=222 |  | 7.3 (0.67) | 7.3 (0.70) | 7.3 (0.66) |
| Age in months round 3, n=214 |  | 11.4 (0.96) | 11.3 (0.99) | 11.5 (0.92) |

Categorical variables are presented as %, continuous variables as mean and standard deviation. <sup>a</sup> An estimate of each household's position within the 2014 Demographic and Health Survey national wealth quintiles using household asset information according to Equity Tool guidelines <sup>b</sup> Improved water source: household piped connections, public standpipes, protected tube wells with a platform, protected dug wells or springs and rainwater collection; <sup>c</sup> Functional handwashing station: equipped with water, detergent, and a pouring device; <sup>d</sup> Access to basic sanitation facilities: flush/pour flush toilets connected to piped sewer systems, septic tanks or pit latrines, pit latrines with slabs (including ventilated pit latrines), and composting toilets not shared with other households; <sup>e</sup> Education: partial/complete primary: up to 5 years of schooling, at least partial secondary: >5 and up to 10 years of schooling or higher education;
Abbreviations: round 1: data collected at child age 0-3 months, round 2: data collected at child age 6-8 months, round 3: data collected at child age 10-13 months

Background characteristics of the FHEED cohort are shown in Table 1. Intervention and control arms were similar regarding household wealth, religion, number of household members, mother’s age at baseline, and mother’s educational attainment. Water, sanitation, and hygiene infrastructure was also largely similar: access to an improved water source was nearly universal (95% control vs. 98% intervention), about half (49% control vs. 60% intervention) had a functional handwashing station, and 32% of control and 45% of intervention households had access to basic latrines. Child age at data and sample collection rounds was, on average, 1.4 months at the first round, 7.3 months at the second, and 11.4 months at the third.

### Enteric pathogens, EED, diarrhea, and child growth: description and intervention effects

Enteric pathogen prevalence in our study population was high (84%) and increased with child age, from 63% at 0-3 months of age to 93% at 10-13 months (Figure 2 and Suppl. Table 2). Children were often infected with more than one pathogen: the median number of pathogens detected was 1 pathogen at 0-3 months, 2 pathogens at 6-8 months, and 3 pathogens at 10-13 months of age. Among the pathogens detected, the most common were *Campylobacter* species, with up to 70% of stool samples positive in 10-13-month-old children, followed by VTEC, with up to 35% positive stool samples in 10-13-month-old children (Figure 2C). The most prevalent viral pathogens were norovirus with 34% and astrovirus with 31% in 6-8-month-old children (Figure 2B). Among protozoan enteric parasites, *Giardia lamblia* and *Cryptosporidium* spp. were the most common, both detected in about 14% of stool samples from 10-13-month-old children (Figure 2D).

**Figure 2:**
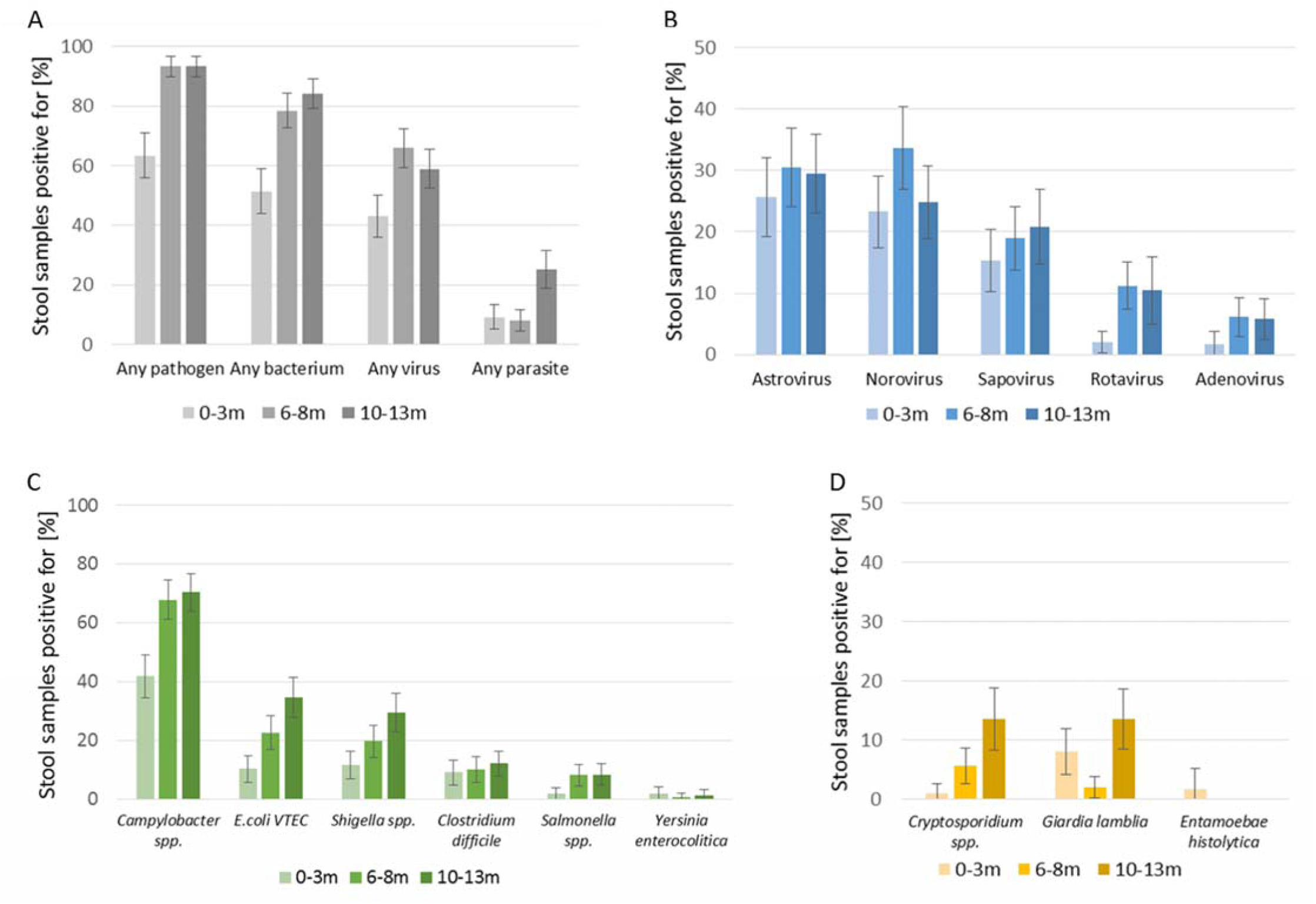
Prevalence of enteric pathogens detected in children’s stool samples by age. Graphs showing the predicted prevalence and 95% confidence intervals of A) pathogen groups, B) viral pathogens, C) bacterial pathogens, and D) parasites in children at 0-3 (light), 6-8 (medium), and 10-13 months of age (dark). Note that axes differ in their maximum value. Prevalences were predicted based on models adjusted for sex of the child, season of sampling, with settlement-level random effects. Total n=636, 0-3 months n=204, 6-8 months n=221, 10-13 months n=211. Abbreviations: *Campylobacter* spp: *Campylobacter jejuni/coli/lari*, E.coli VTEC: verotoxin-producing *Escherichia coli*

There was no evidence that the intervention reduced pathogen prevalence (intervention arm 82%, control arm 86%, OR: 0.75, 95% CI: 0.5-1.2, p=0.25). In fact, intervention children were more likely to have a stool sample positive for sapovirus at 0-3 months of age (21% vs 10% prevalence, OR: 2.3, 95% CI: 1.1-5.2) and for *Giardia lamblia* at 10-13 months of age (19% vs 8%, OR 2.6, CI: 1.1-6.4; Figure 3, Suppl. Table 3 and 4).

**Figure 3:**
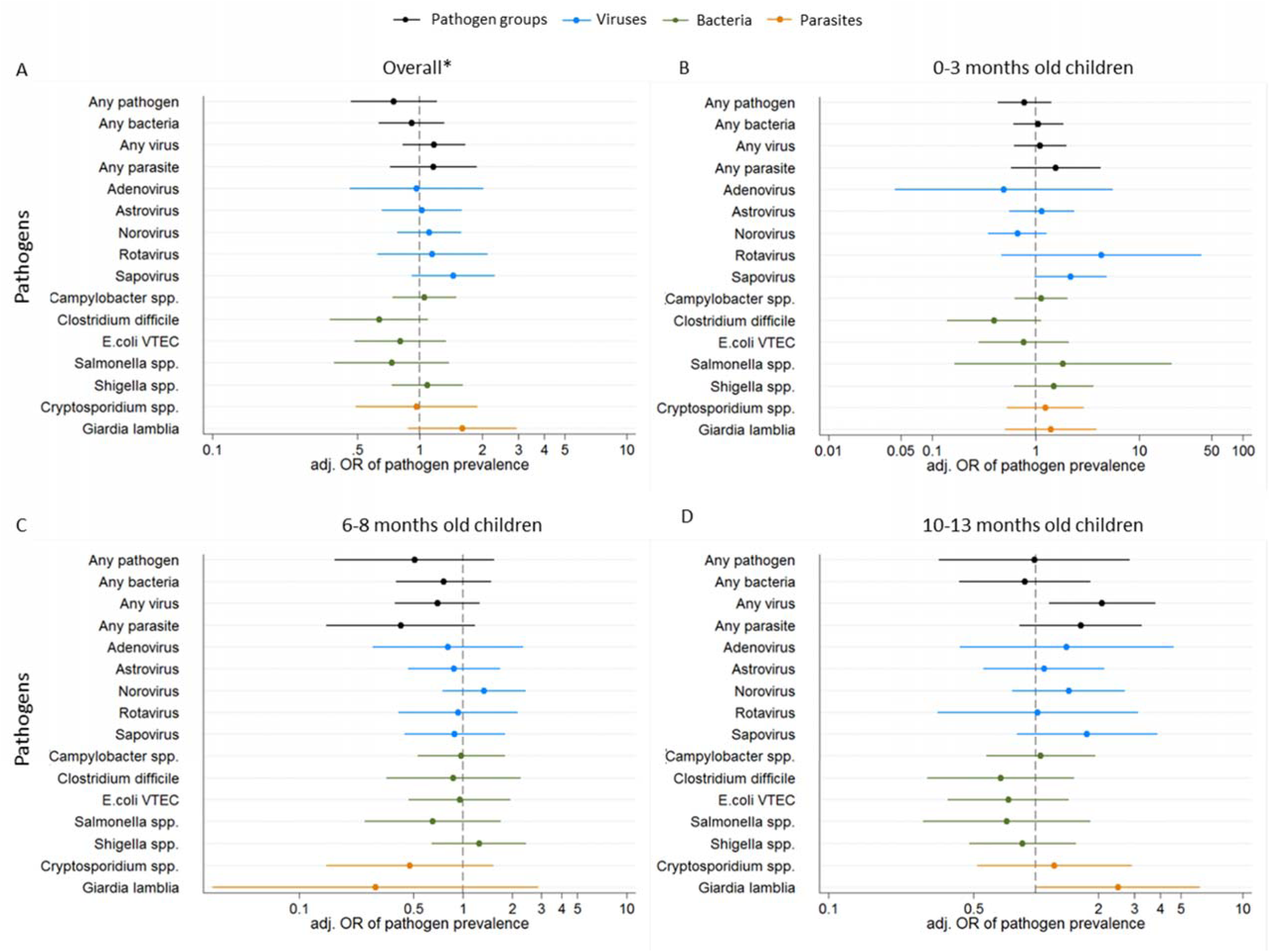
Intervention effect on enteric pathogen prevalence in children overall and by age. Adjusted odds ratios (OR) with 95% confidence intervals of pathogens detected in A) all children, B) children at age 0-3 months, C) children at age 6-8 months, and D) children at age 10-13 months, comparing intervention to control arm. Estimates shown from mixed-effects logistic regression models, adjusted for age (in models indicated with *) and sex of the child, season of sampling, with settlement-level random effects. Total n= 635, Bacteria n=632, Viruses n=628, Parasites n=634 children. Abbreviations: OR: odds ratio, *Campylobacter spp.*: *Campylobacter jejuni/coli/lari*, *E.coli* VTEC: verotoxin-producing *Escherichia coli*

Mean concentrations of EED biomarkers and diarrhea prevalence are presented in Supplementary Table 5. While NEO concentrations showed an increasing trend over the first year of age, there was no clear trend for MPO or AAT. Pairwise correlations between the three biomarkers were rather low in all age groups. The highest correlation was observed between MPO and AAT across ages, with correlation coefficients ranging from 0.34 to 0.48 (Suppl. Table 6). About 5% of mothers reported that their child was sick with diarrhea within the last 7 days, with the highest prevalence (7.6%) at child age 6-8 months. There was no evidence for an intervention effect in the FHEED cohort (consistent with the previously published results for the larger study) on 7-day diarrhea prevalence nor on EED biomarkers (Suppl. Table 7).

Anthropometric measures are presented in Supplementary table 8. Children’s weight was measured over their first year of life, and average weight-for-age z-scores did not change substantially over that period of time, with the lowest weight-for-age z-score being measured at endline (−1.33 mean z-score), when children were between 10-18 months old. Average length-for-age z-scores were comparable at birth and endline (−1.7 at birth, −1.59 at endline), while average weight-for-height z-scores were lower at endline (0.13 at birth, −0.73 at endline).

### Associations between enteric pathogen infection, biomarkers of EED, and child growth

1. Enteric infections and EED biomarkers: In pooled measurements of all age groups, we found that children infected with any parasite had higher MPO concentrations. For single pathogens, the presence of *Giardia lamblia* or *Shigella* spp. was associated with higher MPO concentrations, while children positive for sapovirus had higher AAT concentrations and children positive for *Salmonella* spp. had lower NEO concentrations (Figure 4 and Suppl. Table 9).
2. Enteric infections and growth outcomes: Children who had a parasitic infection at two or more time points over their first year of life had 0.78 lower LAZ and 0.61 lower WAZ at study endline, while the presence of a viral infection at all three time points was associated with 0.68 lower WHZ (Figure 5 and Suppl. Table 10), adjusted for other pathogen groups. Looking at parasitic pathogens (while adjusting for all other pathogens), infection with *Giardia lamblia* at two or more time points was associated with 0.82 lower LAZ, while children positive for *Cryptosporidium* spp. had 0.40 lower WHZ and 0.36 lower WAZ (there was no child infected with *Cryptosporidium* spp. at more than one time point). For viruses, children with a rotavirus infection at two or more time points had 1.97 lower WHZ and 1.56 lower WAZ. Sapovirus infection at two or more time points was associated with 0.55 lower WHZ and 0.54 lower WAZ. There was marginal evidence that infections with bacterial pathogens at any time point were linked to lower LAZ at endline (−0.53, 95% CI - 1.13 to 0.06, p=0.08) and there was no dose-response. In addition to associations between enteric pathogen infections and growth at endline, we examined associations of enteric pathogen infections and WAZ at the time of sampling. Children infected with any parasite had 0.18 lower WAZ, while bacterial or viral infections showed no association (Suppl. Table 11). Analyzing single pathogens, we observed 0.25 lower WAZ in children infected with *Giardia lamblia*, and a marginal association with norovirus infection, with a 0.12 lower WAZ.
3. Biomarkers of EED and growth outcomes: We found no associations between MPO, AAT, or NEO concentrations, averaged over the first year of life, with child growth outcomes at endline (Suppl. Table 12).

## Discussion

This study observed high overall enteric pathogen prevalence in a cohort of rural Bangladeshi infants over their first year of life, despite low caregiver-reported diarrhea, and relatively high levels of EED biomarkers. The combined homestead food production and food hygiene intervention, evaluated in a cluster-randomized trial, did not reduce pathogen prevalence. We observed several associations between enteric pathogens infections, elevated EED markers, and poorer growth outcomes.

**Figure 4:**
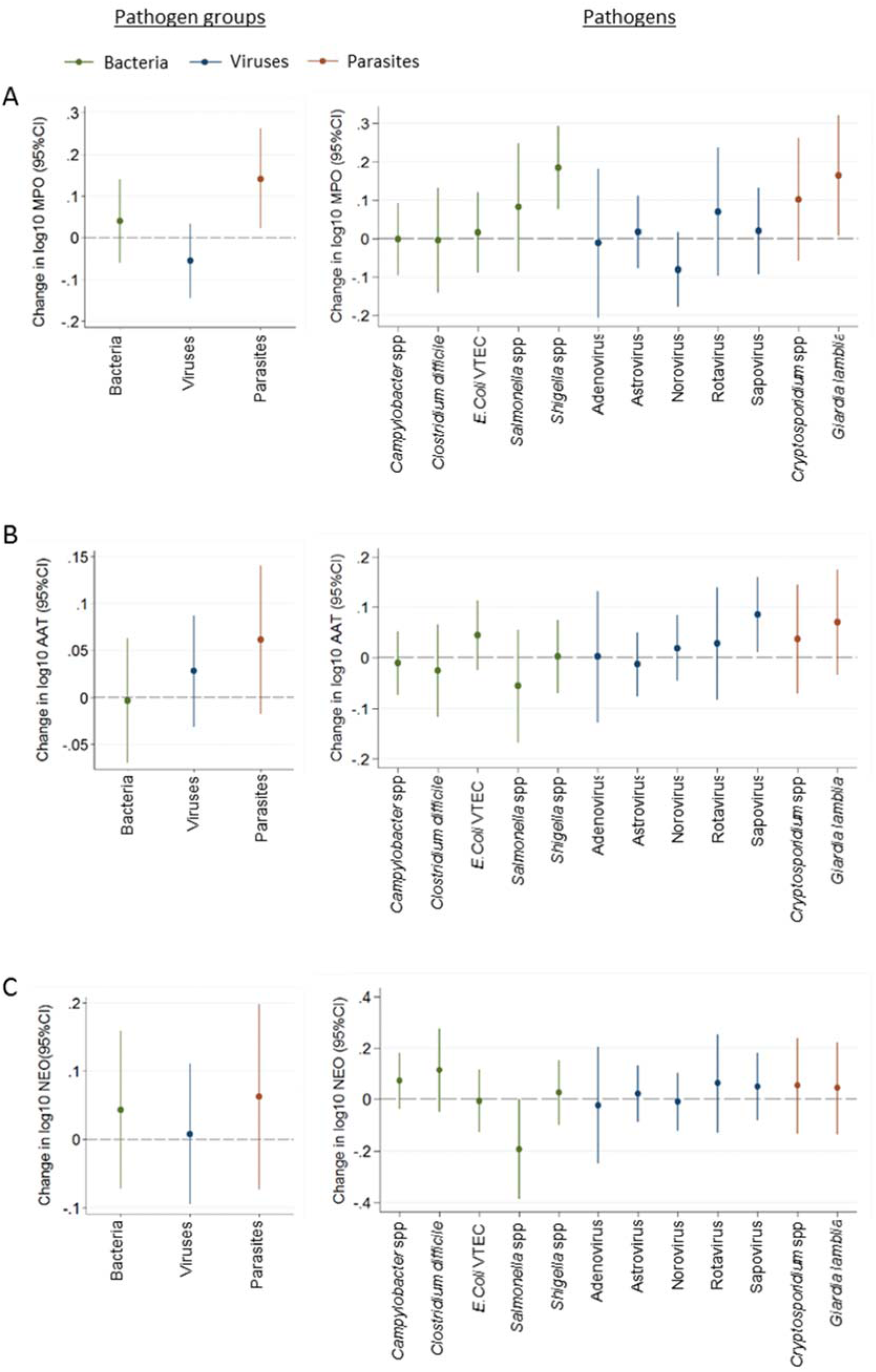
Associations of enteric pathogens with EED biomarkers. Associations shown for pathogen groups (left panel) or single pathogens (right panel) with A) MPO, B) AAT, and C) NEO. Estimates are from mixed-effects linear regression models adjusted for child age and sex, and season of sampling, with settlement- and child-level random effects. For pathogen groups: MPO n= 559, AAT n= 563, NEO n= 547; for single pathogens: MPO n= 552, AAT n= 555, NEO n= 540. Abbreviations: MPO: Myeloperoxidase, AAT: alpha-1-Antitrypsin, NEO: Neopterin, Campylobacter spp.: *Campylobacter jejuni/coli/lari, E.coli* VTEC: verotoxin-producing *Escherichia coli*

**Figure 5:**
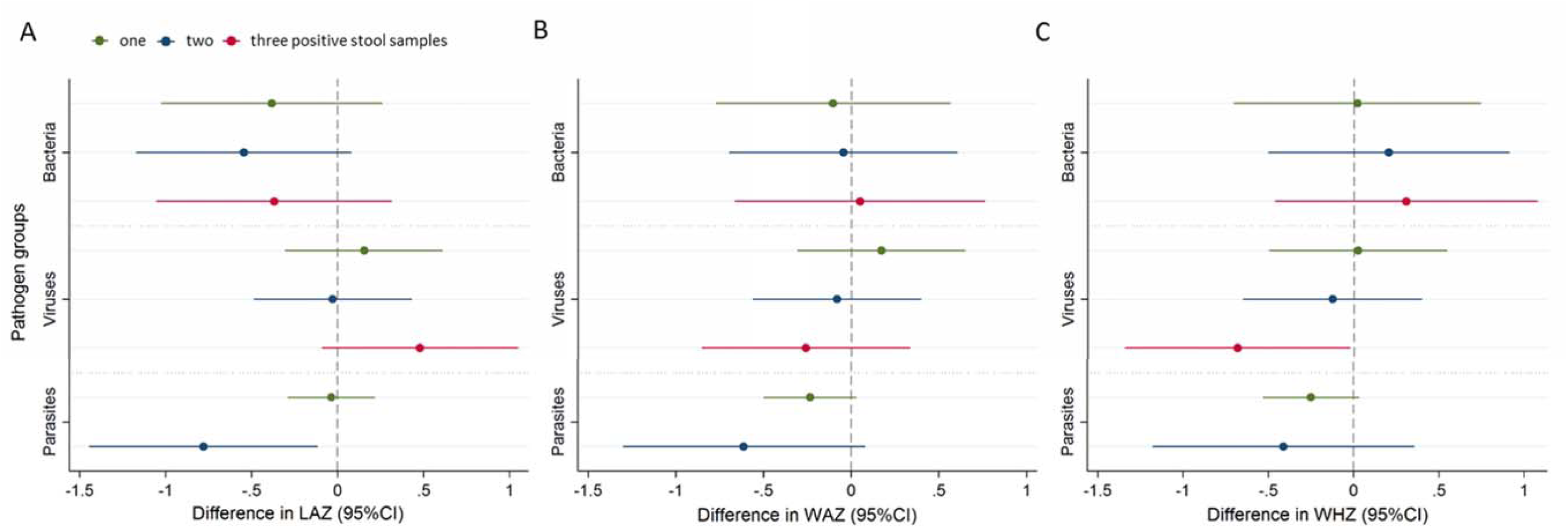
Associations of enteric pathogens with anthropometric measures at study endline. Difference in a) LAZ, b) WAZ, c) WHZ for one (green), two (blue), or three (red) positive stool samples compared to no exposure in all three sampling rounds. For parasite infections, the blue color estimate stands for two or three rounds of infection. Estimates are from mixed-effects linear regression models adjusted for child age and sex, number of rounds sampled, length-for-age (weight-for-age or weight-for-height, respectively) z-score at birth, maternal height, and intervention allocation, with settlement-level random effects. LAZ n= 228, WAZ n=228, WHZ n= 209 children. Abbreviations: LAZ – length-for-age z-score, WAZ – weight-for-age z-score, WHZ – weight-for-height z-score

Pathogen prevalence was over 80%, increased with child age, and most children were infected with more than one pathogen, similar to what was found in previous studies in comparable LMIC settings (9, 10, 14, 30, 41, 42). Although prevalences of specific enteric pathogens varied widely, infections with bacterial pathogens were generally more frequent than viral or parasitic infections, in line with other studies (14, 30, 42). Despite the high pathogen prevalence, on average, only about 5% of caregivers reported diarrhea. In combination with the comparably high levels of EED biomarkers, this indicates widespread asymptomatic enteric infections with ongoing intestinal inflammation in most of the infants in our study, a finding which also has been observed in studies of young children across Asia, Sub-Saharan Africa, and South America (9, 10).

The nutrition and food hygiene intervention had no effect on enteric pathogen prevalence in the FHEED cohort children. This is in line with no improvement observed in diarrhea prevalence and EED biomarkers in the larger FHEED and FAARM study (32, 43). Other recent large trials evaluating combined WASH and nutrition or community-wide water or sanitation interventions have also shown no consistent effects on enteric pathogens. The WASH intervention arms in the WASH Benefits trial in Bangladesh decreased the prevalence of enteric viruses, but not bacteria and parasites (42), while the WASH intervention in the SHINE trial in Zimbabwe decreased the number of detectable parasites, but had no effect on bacteria, viruses, or the prevalence of single pathogens (30). A point-of-use water treatment intervention in South Africa, as well as an urban sanitation intervention in Mozambique, both showed no effect on enteric pathogen prevalence (23, 44). This indicates that our intervention and the other abovementioned WASH interventions were not able to reduce children’s exposure to enteric pathogens at all, or not sufficiently, to achieve a reduction in infections and so could not reduce diarrhea or improve health outcomes like growth.

In the observational part of this study, we found associations between certain enteric pathogens, biomarkers of EED, and growth outcomes. For bacterial pathogens, only *Shigella* was associated with EED biomarkers and growth, while there were no associations between any of the other bacterial pathogens with either EED or children’s growth outcomes at study endline. *Shigella* infection was associated with increased MPO concentrations, in line with previous studies (11, 45). *Shigella* infection over the first year of life was not associated with growth outcomes at study endline, but concurrent *Shigella* infection was associated with lower weight-for-age in cross-sectional analyses. This is somewhat in contrast to findings from other studies that report an association of *Shigella* (21, 41) or other bacterial pathogens (19, 21, 46–49) with reduced height-for-age. These findings, however, were not necessarily consistent across different study settings or in the presence and strength of the observed associations. Other factors, like the age of the child at the time of infection (19), co-infection with other pathogens (19), or complementary food intake and dietary quality (12), are likely to influence the observed effects of a pathogen on child growth.

Parasite infections, mainly with *Giardia*, were associated with increased levels of MPO, as well as decreased linear growth in young children. This is similar to previous studies showing an association of *Giardia* with higher fecal MPO concentrations (18), but also with other biomarkers of epithelial damage and repair, and intestinal or systemic inflammation (19, 48, 50). Associations between *Giardia* infection and lower LAZ in children have also been reported in a number of studies (9, 11, 19, 21, 48, 50–52). However, the mechanistic pathway underlying *Giardia*-associated growth impairment is not fully understood, and recent studies suggest that it is not mediated by pathways of intestinal or systemic inflammation (11) but rather by disruption of nutrient metabolism (53). In our study, even though there was some evidence for an association between *Giardia* infection and increased MPO, and between *Giardia* and decreased LAZ, we did not observe an association between increased MPO levels and decreased LAZ in children, supporting the hypothesis that *Giardia*-associated growth impairment is not mediated through MPO.

Parasitic infections with *Cryptosporidium* in the first year of life showed no clear association with LAZ, but rather with WHZ and WAZ of children at study endline, likely caused by mucosal damage and an inflammatory immune response (54). This is similar to findings from Das et al., reporting that asymptomatic *Cryptosporidium* infections were associated with WHZ and WAZ in young children (51), while symptomatic infections were associated with LAZ (20, 51). Other studies have also found associations between *Cryptosporidium* and LAZ in asymptomatic children, though the evidence was mostly weak (19, 48, 55). In the present study, we were unable to separately investigate symptomatic children due to the low prevalence of diarrhea; only 33 episodes of diarrhea were reported, four of which were concurrent with *Cryptosporidium* infection.

Repeated viral infections during the first year of life, especially with rotavirus but also with sapovirus, were associated with reduced ponderal growth (WLZ and WAZ) of children at study endline. Children with sapovirus infections also showed increased levels of AAT. However, as AAT levels were not associated with child anthropometric parameters in our study, it seems unlikely that the effect of sapovirus (or rotavirus) on ponderal growth was mediated through AAT. An effect on WHZ through symptomatic diarrhea is also rather unlikely in our setting as only very few of the children positive for rotavirus or sapovirus were reported to be sick with diarrhea in the last 7 days. Findings from the Global Enteric Multicenter Study (GEMS) also reported an association between rotavirus and sapovirus as well as norovirus and adenovirus with reduced WHZ and WAZ measures, but less so for LAZ, in asymptomatic children under one year of age (56). Berendes et al. also found a weak association between adenovirus and WHZ in children under 5 years old (9), while two other studies reported associations of norovirus with LAZ (21, 48). While the mechanistic pathways of viral growth retardation in the absence of symptomatic diarrhea remain unclear and require further investigation, our study and the existing evidence show a clear influence of asymptomatic viral infections on early childhood growth.

It may not be too surprising that most of the evaluated WASH and food hygiene interventions have not shown more promising effects on child diarrhea and growth outcomes given the wide range of enteric pathogens found in infants’ guts in this and other studies. Fecal-oral transmission of these pathogens can occur via many different environmental routes, such as water, food, animal contact, or household environment and soil (57, 58). Ours as well as other WASH interventions aimed to prevent the transmission of pathogens via one or a few of these routes, which may have reduced pathogen exposure to some extent, but maintained the risk of contamination or recontamination via other exposure pathways (58). Transformative WASH interventions, which are able to drastically reduce fecal pathogen exposure in children’s home environments from various sources, as well as at the community level, might be needed to ensure a substantial improvement in child health (31, 59, 60).

Our study has several strengths and limitations. The FHEED cohort was embedded in a large cluster-randomized controlled trial. We could make use of the cluster-randomized design and the timely information collected from newborn children through FAARM’s birth surveillance to establish a birth cohort with repeated sampling. This allowed us to study pathogen exposure and biomarkers of EED in children over their first year of life and longer-term consequences on child growth, as well as to examine the intervention impact on pathogens, in addition to biomarkers of EED and diarrhea (both already studied in the larger trial) in more detail. Also, the rich data collected in FAARM provided us with information on important covariates. For pathogen analysis, we tested for 14 of the most common enteric pathogens, which is a large number but still only a subset of all enteric pathogens and therefore provides an incomplete picture of current enteric infection. Also, we cannot make conclusions about whether the pathogens detected in the fecal samples represent a clinical or asymptomatic infection, nor about their intensity and duration, as some of these pathogens are shed for a long time after an active infection. Although we were able to repeatedly measure markers of EED and pathogen exposure in children over their first year of life, financial and organizational constraints only allowed us to take weight, but not length, measures each time we collected a fecal sample, except for at birth (at less than 3 days of age) and FAARM study endline (between ages 12 and 18 months). Therefore, we were not able to look at growth velocity at later ages or potential catch-up growth. Furthermore, our sample size was too small to model more complex pathways to investigate mechanistic effects, for example, a mediation analysis of how much a pathogen’s effect on growth was mediated through biomarkers of EED. We also did not examine the role of the fecal microbiota and its association with enteric infection and inflammation in child growth. However, fecal samples were subjected to metagenomics analyses, and the composition and function of the microbiome and its association with intestinal inflammation and child anthropometric outcomes will be reported in a forthcoming study. Finally, we conducted this study in the FAARM study population in two rural sub-districts of Habiganj District, in Sylhet Division, Bangladesh, which is not nationally representative. Nevertheless, the household environment resembles typical rural areas in Bangladesh and it is thus likely that our results are relevant for places with similar characteristics across the country and other South Asian countries.

## Conclusions

In conclusion, we observed a high burden of enteric pathogens in our study population of infants, despite a low reported prevalence of diarrhea. The combined homestead food production and food hygiene intervention did not result in a reduction in pathogen prevalence. In line with analyses of the larger FAARM and FHEED cohorts (32, 43), we also observed no intervention effect on biomarkers of EED or diarrhea prevalence in the cohort. Some bacterial pathogens and protozoan parasites were found to be associated with EED biomarkers. While parasites were mainly linked to reduced linear growth, viral pathogens showed associations with reduced ponderal growth. Given the range of enteric pathogens associated with child growth and biomarkers of EED and their various exposure pathways, transformative WASH interventions that address multiple sources of pathogen exposure at various levels, including at the community and household levels, are likely necessary to effectively reduce enteric pathogen burden in children’s home environments and to meaningfully improve child health.

## Author’s contributions

SG is the principal investigator of the FAARM trial and FHEED study. SG, AAM, and TNH conceptualized and designed the FHEED study and acquired funding. AAM, SS, and TNH developed data collection tools with guidance from ASW, SG, and other members of the study team. SS and TNH managed the FHEED field team in Bangladesh and supervised data collection. JLW led the data management of the FAARM trial, including the FHEED data, and prepared the initial datasets for this study. DN provided reagents, materials, and other laboratory resources. AAM oversaw laboratory analyses, analyzed the data, prepared and interpreted the results, and wrote the first and subsequent drafts of this article. NJL supported the data analysis and interpretation of results. MR provided general supervisory and management support to the FHEED team in Bangladesh. AAM wrote the original draft of the manuscript and all authors critically reviewed the manuscript in several steps and approved the final version.

## Funding source and their role

The German Federal Ministry of Education and Research is the primary funder for the FAARM trial (grant no. 01ER1201). The FHEED study, nested within FAARM, is supported financially by a project grant from Deutsche Forschungsgemeinschaft (German Research Foundation, project no. 413269709). FHEED’s research work was further supported by Foundation Fiat Panis. Additional support was provided to Helen Keller International by the Carrefour social responsibility program and other charitable donations for the implementation of the HFP program. Funding organizations had no role in the FAARM trial design, the intervention or its implementation, in training, data collection, analysis, and interpretation of results.

## Data Availability

The data underlying this study will be deposited in a publicly accessible repository upon acceptance. Accession numbers and/or DOIs will be provided at that time.

## Acknowledgements

We thank the FAARM households for their valuable time and participation. We thank our collaboration partners: Helen Keller International and icddr,b in Bangladesh. We are grateful to the survey staff and supervisors, especially to Saheen Hossein, for their time and effort in sample and data collection; and to Abdul Kader for his support in questionnaire adaptation and training. We also thank Dr. Jesmin Sultana and Dr. Wolf-Peter Schmidt for their valuable feedback and support in the design of the FHEED study, and Sushobhan Sarker for his support in the preparation of data collection, biosampling, training, and procurement. Furthermore, we thank Alicia Schäfer, Simran Rastogi and Nikolaus Gössl for their valuable assistance with laboratory analyses, and the Medical Microbiology Laboratory staff, Heidelberg University Hospital, especially Aline Sähr and Sandra Förmer for their kind support. We are also grateful to Dr. Tobias Kurth for his valuable guidance on data analysis.

## Supplementary Tables - Captions

**Suppl. Table 1: Model specifications including covariate adjustment and random effects.** ^a^ Pathogen groups: any pathogen, any bacterium, any virus, any parasite (that was measured); ^b^ Single pathogens: Adenovirus, Astrovirus, Norovirus, Rotavirus, Sapovirus, Campylobacter spp., Clostridium difficile, E.coli VTEC, Salmonella spp., Shigella spp., Yersinia enterocolitica, Cryptosporidium spp., Entaboebae histolytica, Giardia lamblia; ^c^ in case mixed-effects logistic regression models did not converge, we used cluster-robust standard errors with clustering at settlement level, this was the case for: Adenovirus, Rotavirus, Shigella spp., Salmonella spp., Yersinia enterocolitica, Entamoebae histolytica; ^d^at the time of the FHEED cohort survey; ^e^ at FAARM endline survey.

**Suppl. Table 2: Prevalence of enteric pathogens detected in children’s stool samples by age.** 0-3 months n= 204, 6-8 months n= 221, 10-13 months n= 211. Prevalences were predicted based on models adjusted for sex of the child, season of sampling, with settlement-level random effects; Dash (-): not detected. Abbreviations: Prev.: predicted prevalence, CI: confidence interval, *Campylobacter spp.*: *Campylobacter jejuni/coli/lari*, E.coli VTEC: verotoxin-producing *Escherichia coli*

**Suppl. Table 3: Enteric pathogen prevalence by intervention arm in children overall and by age.** Total n= 635, Bacteria n=632, Viruses n=628, Parasites n=634. Prevalences were predicted based on models adjusted for age (in models indicated with *) and sex of the child, season of sampling, with settlement-level random effects; Abbreviations: Interv.: Intervention, Pred. prevalence: predicted prevalence, Pred. number: predicted number, *Campylobacter* spp.: *Campylobacter jejuni/coli/lari*, E.coli VTEC: verotoxin-producing *Escherichia coli*

**Suppl. Table 4: Intervention effect on enteric pathogen prevalence in children overall and by age.** Total n= 635, Bacteria n=632, Viruses n=628, Parasites n=634. Models were adjusted for age (in models indicated with *) and sex of the child, season of sampling, with settlement-level random effects. Abbreviations: OR: odds ratio, CI: confidence interval, p: p-value, *C. jejuni/coli/lari*: *Campylobacter jejuni/coli/lari*, VTEC: verotoxin-producing *Escherichia coli*

**Suppl. Table 5: Concentration of EED biomarkers and diarrhea period prevalence by child age.** Total n: MPO n=565, AAT n=569, NEO n=552, diarrhea n=641. The table shows predicted concentrations and 95% CI of markers of environmental enteric dysfunction in stool, and predicted prevalence and 95% CI of diarrhea 7-day period prevalence in children at 0-3 months, 6-8 months, and 10-13 months of age. Estimates were predicted based on models adjusted for sex of child, season of sampling, with child-level and settlement-level random effects. * n is lower because in 44 children at age 0-3 months we were not able to get any or only very few native sample and could therefore not perform the biomarker analyses. Abbreviations: MPO: Myeloperoxidase; AAT: Alpha-1-Antitrypsin; NEO: Neopterin; Pred. conc.: predicted concentrations; Pred. prev.: predicted prevalence; CI: confidence interval.

**Suppl. Table 6: Pairwise correlation of EED biomarkers, by child age.** Number of observations included in the single comparisons are given in each cell; Abbreviations: MPO: Myeloperoxidase, AAT: Alpha-1-Antitrypsin, NEO: Neopterin, r: correlation coefficient.

**Suppl. Table 7: Intervention effect on EED biomarkers and diarrhea prevalence by child age.** MPO n=565, AAT n=569, NEO n=552, Diarrhea n=640. From mixed-effects linear (for MPO, AAT, NEO concentration) or logistic (for diarrhea prevalence) regression models, adjusted for child sex and season of sampling, with settlement-level and child-level random effects. Abbreviations: MPO: Myeloperoxidase, AAT: alpha-1-Antitrypsin, NEO: Neopterin, Pred. conc.: predicted log10 concentration, Pred. prevalence: predicted prevalence, Coef.: regression coefficient, OR: odds ratio, CI: confidence interval

**Suppl. Table 8: Anthropometric measures of FHEED cohort children at birth, sample collection rounds and study endline.** Birth n=231, Age 0-3months n=113, Age 6-8 months n=222, Age 10-13months n=212, Endline n= 227. ^a^ birth anthropometry measures were taken later than 28 days of age in 7 children and these measures were replaced by average birth anthropometry measures of the cohort. Abbreviations: CI: confidence interval

**Suppl. Table 9: Association of enteric pathogens and EED biomarkers.** For model 1: MPO n= 559, AAT n= 563, NEO n= 547, for model 2: MPO n= 552, AAT n= 555, NEO n= 540. Mixed-effects linear regression models were adjusted for child age and sex, and season of sampling, with settlement- and child-level random effects. Abbreviations: MPO: Myeloperoxidase, AAT: Alpha-1-Antitrypsin, NEO: Neopterin, Pred. conc.: predicted log10 concentration, Coef.: regression coefficient, CI: confidence interval, p: p-value, *Campylobacter* spp.: *Campylobacter jejuni/coli/lari*, E.coli VTEC: verotoxin-producing *Escherichia coli*

**Suppl. Table 10: Association between enteric pathogen exposure in the first year of life and child growth at study endline.** For both models: LAZ n= 228, WAZ n=228, WHZ n= 209. Mixed-effects linear regression models were adjusted for child age and sex, number of rounds sampled, length-for-age (weight-for-age or weight-for-height, respectively) z-score at birth, maternal height, and intervention allocation, with settlement-level random effects; birth anthropometry measures that were taken later than 28 days of age were replaced by average birth anthropometry measures of the cohort. Dash(-): not detected. Abbreviations: Coef.: regression coefficient, 95% CI: 95% confidence interval, p: p-value, *Campylobacter* spp*: Campylobacter jejuni/coli/lari*, E.coli VTEC: verotoxin-producing *Escherichia coli*

**Suppl. Table 11: Association of enteric pathogen exposure and child weight-for-age z-score at the time of sampling.** Sample sizes are n= 543 (model 1), n= 535 (model 2). Mixed-effects linear regression models were adjusted for child age and sex, weight-for-age z-score at birth, maternal height, and intervention allocation, with settlement- and child-level random effects; birth anthropometry measures that were taken later than 28 days of age were replaced by average birth anthropometry measures of the cohort. Abbreviations: Coef.: regression coefficient, 95% CI: 95% confidence interval, p: p-value, *Campylobacter* spp.: *Campylobacter jejuni/coli/lari*, E.coli VTEC: verotoxin-producing *Escherichia coli*

**Suppl. Table 12: Association of EED biomarkers averaged over the first year of life and anthropometric measures at study endline.** LAZ n= 220, WAZ n=220, WHZ n= 201. EED biomarker concentration was averaged over the three rounds of sample collection. Mixed-effects linear regression models were adjusted for child age and sex, length-for-age, weight-for-age, or weight-for-height z-score at birth, respectively, rounds of samples taken, maternal height, and intervention allocation, with settlement-level random effects; birth anthropometry measures that were taken later than 28 days of age were replaced by average birth anthropometry measures of the cohort. Abbreviations: Coef.: regression coefficient, 95% CI: 95% confidence interval, p: p-value

